# Religious beliefs and practices, political orientation, and distrust in healthcare predict attitudes toward mRNA vaccines in the United States

**DOI:** 10.64898/2026.04.06.26350267

**Authors:** Erin D. Solomon, Eu Gene Chin, Kari Baldwin, Lauren L. Baker, James M. DuBois

## Abstract

Religion has contributed to societal divides regarding COVID-19 mRNA vaccines. In this study, we conducted a secondary analysis of a survey of U.S. adults (*N*=4939) focused on how religious affiliations, beliefs, and practices impact attitudes toward genetic and genomic activities, one of which was mRNA vaccines. The dataset included large samples of participants from six religious groups in the U.S. (Black Protestant, Catholic, Evangelical Protestant, Jewish, Mainline Protestant, and Muslim), as well as individuals who were atheist, agnostic, or spiritual. ANCOVA results indicated that Evangelical Protestant participants showed significantly less support for mRNA vaccines than other groups, while atheist participants were the most supportive. Muslim participants had the highest concerns, whereas atheist participants had the lowest. Regression analyses indicated the strongest predictors of support for mRNA vaccines were more spiritual community support for community health, followed by higher acceptance of evolution, more liberal political orientation, less distrust toward the healthcare system, higher frequency of attending religious activities, higher income, lower fundamentalist religious beliefs, and more spiritual community support for liberal reproductive and end of life views. The strongest predictors of concerns about mRNA vaccines were more distrust toward the healthcare system and more conservative political orientation, followed by less spiritual community support for community health, stronger beliefs about God in the body, more fundamentalist religious beliefs, and lower knowledge of genetics. The large sample size, and examination of a broad array of religious variables alongside distrust and political orientation offer new insights. These findings add to the literature on the culture wars surrounding mRNA vaccines, and can perhaps aid in future efforts to build trust and relationships between public health and religious communities.

## Introduction

In a recent article, Gostin and Wetter lamented that evidence-based practices that save lives—including specifically Covid-19 vaccines—are caught in the cross hairs of culture wars.(1) They characterize these wars as political, reflecting partisan divides. Davison Hunter, the sociologist who coined the term “culture war” believed they arose primary between conservative religious factions and secular society.(2) Most Covid-19 vaccines are mRNA vaccines, and were the first mRNA vaccines to be administered widely.(3-6) In this paper, we draw upon data from a survey of nearly 5,000 adults in the U.S. to explore how religion, politics, distrust, and other demographic variables influence attitudes toward mRNA vaccines.(7)

The modest body of literature on religion and vaccines is primarily theoretical rather than empirical. One recent framework suggests that religious factors influencing vaccine attitudes generally fit into a few types.(8) One type involves an individual or faith group’s religious worldview. For example, some religions rely on faith and prayer for healing, rather than medical interventions like vaccines, because they view disease as being caused by sin or evil or they emphasize trust in God for healing.(8-10) Another type centers around the idea of purity. Some religions believe that vaccines are impure because they are unnatural or the ingredients used to manufacture vaccines are impure, and as such are unfit for the human body.(8, 11, 12)

Morality is another religious factor influencing vaccine attitudes. Some religions view vaccines as problematic because of the use of gelatin in the vaccine and vaccine development, which can be derived from sacred animals, or animals that were not slaughtered in accordance with religious practices.(8, 13, 14) Beliefs about the sanctity of life for embryos and fetuses are rooted in moral concerns about the use of embryonic or fetal tissue in vaccine development.(8, 13) However, though religious beliefs such as these are cited as concerns about vaccines, prior research has also suggested that at least in some cases, religious concerns about vaccines actually reflected concerns about vaccine safety or personal beliefs.(14)

Religions have been described as “moral communities,” in which communal beliefs can significantly shape individual behavior.(15) When a religion indicates that vaccines are morally problematic or should not be used, it demonstrates how an individual-level decision can be influenced by the broader religious context. Additionally, much of our moral decision making is based on moral intuition or intuitive ethics—a fast, heuristic, outside of conscious awareness type of thinking that involves quick feelings of either approval or disapproval in relation to moral situations (i.e., a “gut reaction”).(16, 17) This type of thinking reinforces how ingrained religious beliefs and communal values can unconsciously shape individual decision making.(16, 17) Thus, adopting a theoretical approach to understanding attitudes toward mRNA vaccines may fail to explain real-world attitudes and behaviors.

Religious affiliation and religious beliefs appear to play a major role in shaping attitudes toward vaccines.(18-22) Prior research has shown that individuals and communities higher in religiosity and who frequently attend religious services tend to have lower vaccination rates and lower support for vaccines.(23-26) However, these studies do not explore what specific features of religious engagement might explain vaccine hesitancy. Further, most of the prior literature that examines the influence of religion has focused on specific religious groups and their views of vaccines.(8, 11, 12, 27-32) In rare cases, specific communities such as Orthodox Jews and the Amish, were examined following recent outbreaks of diseases like measles and polio, which occurred due to low vaccination rates.(33-35)

### mRNA Vaccines

Most large empirical studies of Covid-19 mRNA vaccine attitudes have not examined the role of religious affiliation, beliefs, or practices on attitudes or acceptance of vaccines. For example, Iqbal et al report on the Global Listening Project, which conducted an interview-based survey of 70,000 adults from 70 nations on willingness to accept new mRNA Covid vaccines, but never inquired into how religion might influence participant’s willingness.(36) Another global survey of 7,000 adults across 7 nations explored trust and mistrust of new Covid-19 vaccines, but did not inquire into religious variables.(37) In 2022-23, Xu and colleagues reviewed over 541,000 tweets on Twitter related to confidence in mRNA vaccines, but focused solely on sentiments and confidence in the vaccines.(38) In contrast to these studies, Odame et al examined 2,858 Twitter posts using a qualitative interpretative approach, and found that mistrust in vaccine science, politics, and religious attitudes were featured most prominently as reasons for concerns regarding mRNA vaccines.(39) Other research has found that countries with more religious populations tended to have lower COVID-19 vaccination rates.(40) Additionally, some religious groups report higher levels of COVID-19 mRNA vaccination than non-religious individuals (e.g., Church of England, Roman Catholics), while other groups report lower levels (e.g., Muslim, Evangelical Protestant, Pentecostal).(41, 42)

### The Current Study

To summarize, much of the literature on religion and vaccines has been theoretical, and while empirical studies have documented an association between religion and vaccine attitudes, few studies have compared religious groups or explored specific aspects of religion that might predict vaccine attitudes. Having a better understanding of the attitudes of major religious groups and which religious beliefs and practices impact attitudes toward mRNA vaccines can inform public health efforts and has the potential to build public trust. Research focused on building trust in public health generally focuses on building trust among racial minorities,(43, 44) and very little research has focused specifically on building trust among different religious groups. Thus, it is possible that building trust with religious groups may be an unutilized lever for improving vaccination rates and public health. Additionally, because the U.S. public attends religious services three times more than scientists in the U.S., it is important that scientists study the role of religion in public health because they are at risk of being less informed by personal experience.(45)

In the current study, we conducted a secondary analysis of a large sample of U.S. adults.(7) The data was from an online survey of religious affiliation, beliefs, and practices, as well as attitudes toward various genetic and genomic activities, one of which was mRNA vaccines. Analyzing data from a large sample allowed us to 1) compare and contrast various major religions within the U.S. on their attitudes toward mRNA vaccines, as well as 2) examine which specific religious beliefs and practices contribute toward attitudes toward mRNA vaccines.

## Materials and Methods

### Study Design

This study was a secondary analysis of cross-sectional survey data,(46) which is appropriate for exploratory studies aimed at testing differences between groups and building predictive models. The data was originally collected via an online Qualtrics survey battery that consisted of measures assessing participants’ religious and spiritual beliefs and practices, including integration of faith into daily living, religious fundamentalism, views toward creation and evolution, beliefs about God in the body, religious discrimination, beliefs that God controls everything, the healthcare values of their spiritual community, frequency of and time spent in prayer and meditation, and frequency of volunteering in religious group activities.(7, 47-53) The survey also included the Attitudes toward Genomics and Precision Medicine (AGPM) measure, measures of genetic knowledge, healthcare system distrust, and participant demographics.(7, 54-57) Further details on the survey and data collection can be found in a prior publication.(7)

Support for the use of mRNA vaccines was assessed with an AGPM item “*I generally support the use of mRNA vaccines.*” Concerns with mRNA vaccines were assessed using the mean score of responses to two AGPM items: “*I worry that people will be required to get mRNA vaccines*,” and “*I am concerned that mRNA vaccines might change human bodies in ways that are unknown*.” For all three items, response options ranged from 1 (strongly disagree) to 7 (strongly agree).

Because some of our analytical decisions were data-driven, we share some data in our Methods section, while reserving primary findings for the Results section. Further details on the analytical decisions are in the *Supplemental Materials*. The study was approved by the Institutional Review Board (IRB) at Washington University in St. Louis (IRB #202201153).

### Sample Characteristics

The original sample of participants (*N*=4939) were recruited using two panel service companies, Prolific and CloudResearch. From Prolific, participants were recruited who were representative of the U.S. population in terms of age, gender, and race (*n*=2999). From CloudResearch, participants were recruited in stratified samples of at least 300 participants from each of six religious groups: Black Protestant, Catholic, Evangelical Protestant, Jewish, Mainline Protestant, and Muslim (*n*=1940). To qualify for the original study, participants needed to be 18 years or older and located in the U.S. Demographic details can be found in a prior, open-access publication.(7)

### Statistical Analysis

To answer the two research questions, the study was divided into two major analytical sections. The first analytical section was comprised of two Analysis of Covariance (ANCOVA) models: one predicting support for the use of mRNA vaccines, and the other predicting concerns about mRNA vaccines. The second analytical section was comprised of two backward chunkwise elimination model building procedures(58): one predicting support for the use of mRNA vaccines and the other predicting concerns about mRNA vaccines. Tables 1 and 2 depict the bivariate correlations between predictor variables used in the ANCOVAs and backward chunkwise elimination model building procedures with each outcome measure.

**Table 1.** Rank-ordered correlations for predictors of support for mRNA vaccines.

| Rank | Predictor Variable | Support |
| --- | --- | --- |
| 1 | Conservative political orientation | -.40* |
| 2 | Acceptance of evolution | .36* |
| 3 | Spiritual community's support for community health | .34* |
| 4 | Spiritual community's support for liberal reproductive & end of life views | .28* |
| 5 | Fundamentalist religious beliefs | -.25* |
| 6 | Private prayer frequency | -.21* |
| 7 | Distrust towards the health care system | -.20* |
| 8 | Education | .20* |
| 9 | Private prayer time | -.18* |
| 10 | Beliefs about God in the body | -.16* |
| 11 | Belief that God controls everything | -.16* |
| 12 | Household Income | .15* |
| 13 | Integration of religious or spiritual beliefs in daily living | -.13* |
| 14 | Religious discrimination: Felt need to conceal religious identity from others | .10* |
| 15 | Age | -.08* |
| 16 | Meditation frequency | -.07* |
| 17 | Frequency of volunteering within your religious or spiritual group | .06* |
| 18 | Meditation time | -.06* |
| 19 | Health in the last four weeks | -.06* |
| 20 | Religious discrimination: Belief that others discriminate against them for their religion | .01 |
| 21 | Genetic Knowledge Index | -.01 |
| 22 | Attendance frequency in religious or spiritual group activities | .01 |

*Note.* This table only includes scale and ordinal predictors that were used in the ANCOVA and backward chunkwise elimination model building procedure. Pairwise deletion was used for these bivariate correlations, with sample sizes ranging from $N = 3431$ to $N = 4439$ , depending on which pairwise relationship is being analyzed. \* $p < .001$ (2-tailed)

**Table 2.** Rank-ordered correlations for predictors of concerns about use of mRNA vaccines.

| Rank | Predictor Variable | Concerns |
| --- | --- | --- |
| 1 | Conservative political orientation | .46* |
| 2 | Acceptance of evolution | -.35* |
| 3 | Private prayer time | .34* |
| 4 | Fundamentalist religious beliefs | .34* |
| 5 | Belief that God controls everything | .33* |
| 6 | Distrust towards the health care system | .32* |
| 7 | Private prayer frequency | .32* |
| 8 | Beliefs about God in the body | .31* |
| 9 | Integration of religious or spiritual beliefs in daily living | .28* |
| 10 | Genetic Knowledge Index | -.22* |
| 11 | Meditation time | .21* |
| 12 | Meditation frequency | .20* |
| 13 | Attendance frequency in religious or spiritual group activities | .17* |
| 14 | Frequency of volunteering within your religious or spiritual group | .16* |
| 15 | Spiritual community's support for community health | -.16* |
| 16 | Religious discrimination: Belief that others discriminate against them for their religion | .14* |
| 17 | Religious discrimination: Felt need to conceal religious identity from others | .11* |
| 18 | Education | -.08* |
| 19 | Spiritual community's support for liberal reproductive & end of life views | -.08* |
| 20 | Health in the last four weeks | -.06* |
| 21 | Age | .02 |
| 22 | Household Income | .00 |

*Note.* This table only includes scale and ordinal predictors that were used in the ANCOVA and backward chunkwise elimination model building procedure. Pairwise deletion was used for these bivariate correlations, with sample sizes ranging from $N = 3431$ to $N = 4439$ , depending on which pairwise relationship is being analyzed. \* $p < .001$ (2-tailed)

### Analysis of Covariances (ANCOVAs)

The ANCOVAs compared each outcome across the nine religious and non-religious groups, accounting for several covariates. The nine religious and non-religious groups were (1) Black Protestant, (2) Catholic, (3) Evangelical Protestant, (4) Mainline Protestant, (5) Jewish, (6) Muslim, (7) atheist, (8) agnostic, and (9) spiritual but not religious (hereafter “spiritual”). Five hundred participants were not able to be included in the ANCOVA models because they did not belong to one of the nine religious/non-religious groups (e.g., Buddhist, Hindu, Mormon) and these subgroup samples were too small to analyze. Based on a review of prior literature, nine demographic covariates were selected: (1) age, (2) education, (3) household income, (4) conservative political orientation, (5) employment, (6) urban/suburban/rural status, (7) gender, (8) race, and (9) ethnicity. See the *Supplemental Materials* for a description of the ANCOVA assumption checking results.

For the ANCOVA predicting support for use of mRNA vaccines, in preliminary checks, we conducted bivariate correlations and one-way ANOVAs between the covariates and support for mRNA vaccines. Only covariates that had a (1) statistically significant relationship with the outcome and (2) evidenced at least a small effect size (e.g., *r* ≥ .10; partial *η*^2^ ≥ .0.02) were considered for inclusion in the model.(59-61) Three covariates (i.e., education, *p* < .001, *r* = .20; household income, *p* < .001, *r* = .14; and political orientation, *p* < .001, *r* = -.41) met the these criteria. These three covariates were assessed for collinearity; none of them had large effect sizes (e.g., *r* ≥ .50; partial *η*^2^ ≥ .0.26) and thus all were included in the ANCOVA.(59, 60) Additionally, 116 participants were not able to be included when their responses could not be used in statistical analyses, such as “prefer not to answer” or “other” income and education respectively, resulting in a total sample size of *n*=4,323 for this ANCOVA.

Similarly, for the ANCOVA predicting concerns about mRNA vaccines, we again conducted bivariate correlations and one-way ANOVAs between the covariates and the mean of the two concerns about mRNA vaccines items. Of the nine covariates, only one (i.e., conservative political orientation, *p* < .001, *r* = -.46) met the criteria for inclusion in the model. Collinearity was assessed, and conservative political orientation was not strongly related to the religious or non-religious group variable (e.g., *r* ≥ .50; partial *η*^2^ ≥ .0.26), thus it was included as a covariate in the ANCOVA model.(59, 60) The final sample size for the ANCOVA was *n*=4439.

### Model Building: Backward Chunkwise Elimination Procedure

The backward chunkwise elimination procedure was conducted to build two regression models: (1) one to predict support for mRNA vaccines, and (2) one to predict concerns about mRNA vaccines. We identified 28 prospective predictor variables to include in the maximum model, which could be broadly divided into four categories: demographic variables, general covariates, religious group, and religious or spiritual variables.(7) Because the model building procedures involved religious or spiritual variables, participants who reported being atheistic or agnostic (*n* = 1008) were not able to be included in the model building procedures because they did not complete the religious or spiritual survey items. Additionally, participants who could not be categorized into any of the religious or non-religious groups (*n*=500) were excluded from the model building procedures, leaving *n*=3431 participants. Subsequently, this subset of participants was randomly divided into a model building (training) group (*n*=1715) and model reliability testing (holdout) group (*n*=1716). More details about the backward chunkwise elimination procedure, maximum models, and reliability indices are described in the *Supplemental Materials*.(58)

Based on the model building sample, employment was removed from the models because it was deemed to be too collinear with age (*p* < .001, *ε*^2^= .43). Additionally, using the model building sample, we conducted preliminary correlations and ANOVAs between each predictor and each outcome (i.e., support and concerns) to exclude predictors that were either not statistically significant (*p*<.05) or had a small effect size (*r* < .1; *ε*^2^<.01; *R^2^* <.02).(60, 62)

For the model predicting support, we sequentially removed predictors that produced a minimum test statistic *F_p_* that was smaller than *F_CRIT_,* where α = .0023 until the minimum test statistic *F_p_* in the model was larger than *F_CRIT_*, leaving us with nine predictors.(63) These predictors were then subjected to a single variable backward elimination procedure using the same inclusion criterion (α = .0023). With the exception of frequency of volunteering, eight of the nine variables met the criterion for inclusion. Thus, these eight predictors were entered into a hierarchical regression, arranged according to previously mentioned conceptual categories.(7)

Similarly, for the model predicting concerns, we sequentially removed predictors that produced a minimum test statistic *F_p_* that was smaller than *F_CRIT_,* where α = .0023 until the minimum test statistic *F_p_* in the model was larger than *F_CRIT_*, leaving us with seven predictors.(63) These predictors were then subjected to a single variable backward elimination procedure using the same inclusion criterion (α = .0023). With the exception of their perception of healthcare values of their spiritual community relating to reproductive and end of life issues, six of the seven variables met the criterion for inclusion. Thus, these six predictors were entered into a hierarchical regression, arranged according to previously mentioned conceptual categories.

## Results

### Dispersion of Mean Support Scores

Figure 1 presents the distribution of participants’ Likert ratings for level of support for the use of mRNA vaccines with higher scores indicating higher levels of support. The mean score of 5.48 (*SD*=1.70) roughly corresponds to a response between “somewhat agree” (a score of 5) and “agree” (a score of 6). Only 24.35% of respondents had mean ratings at or below neutral.

**Figure 1.**
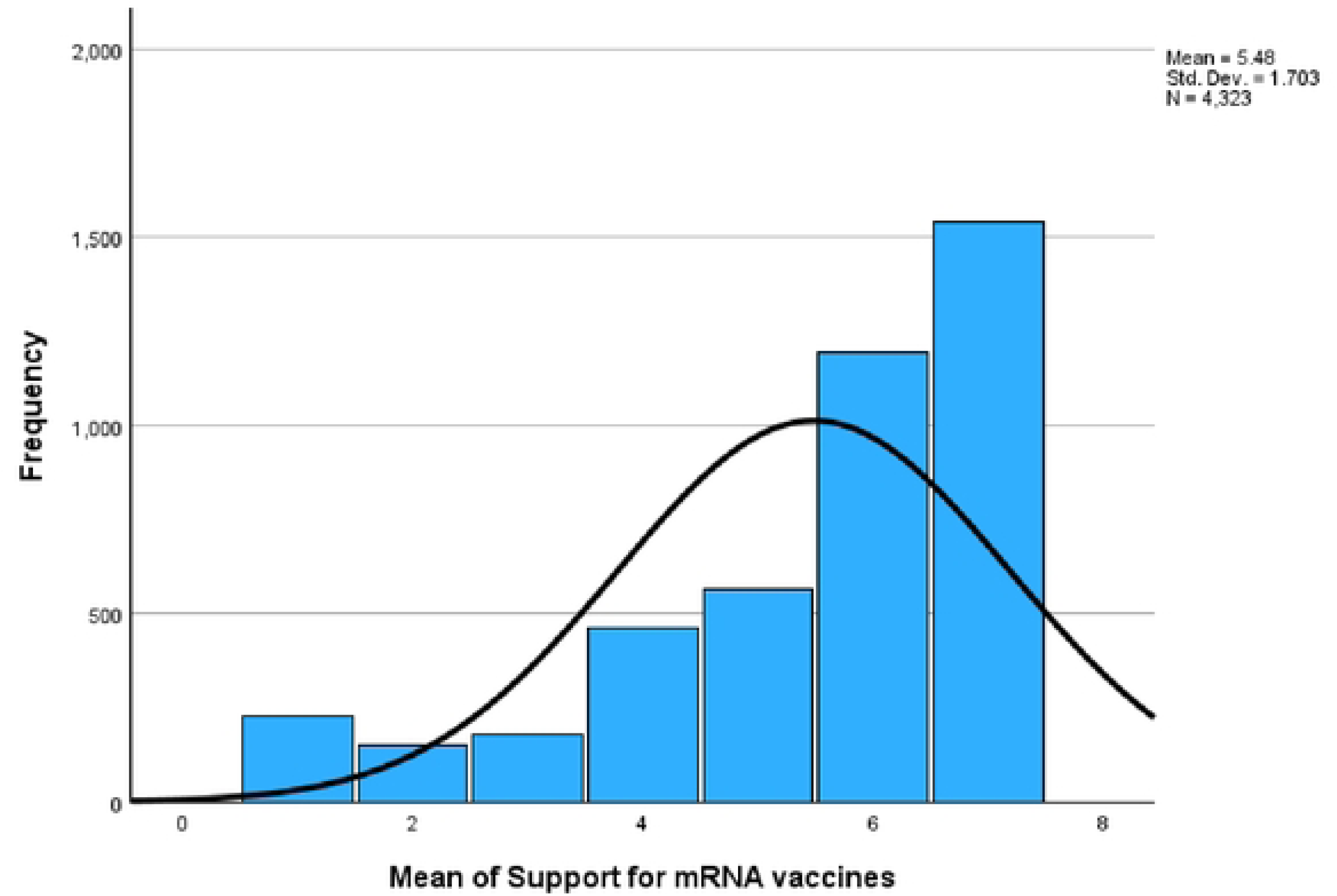
Dispersion of support for mRNA vaccines.

Figure 2 presents the distribution of participants’ mean level of concerns about the use of mRNA vaccines with higher scores indicating higher levels of concern. The mean score of 3.70 (*SD*=1.89) roughly corresponds to a response of “neither agree nor disagree” (a score of 4). The majority of respondents had mean ratings at or below neutral (61.30%).

**Figure 2.**
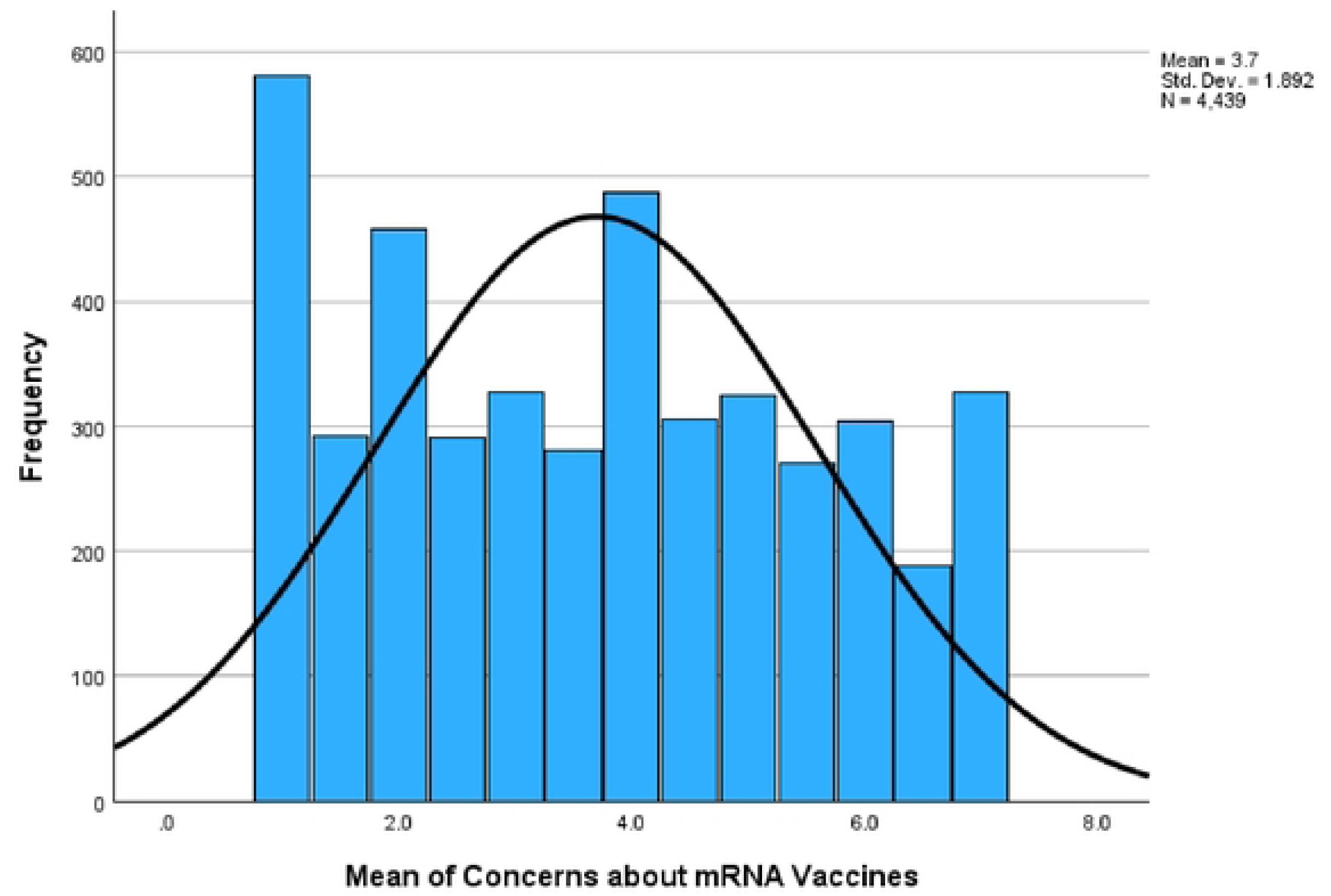
Dispersion of concerns about mRNA vaccines.

### Analysis 1: ANCOVA comparing support for mRNA vaccines among religious and non-religious groups

The ANCOVA results indicated medium sized differences (partial *η*^2^ = .019) across the religious and non-religious groups in terms of support for mRNA vaccines, *F (*8,4323) = 43.23, *p* < .001). Pairwise comparison between groups suggested that the atheist group had a significantly higher mean for support for use of mRNA vaccines compared to all other groups (Δ *M* ranged from 0.35 to 0.74, with all *p*s < .05), except for the agnostic group (Δ*M* = 0.18, *p* = .92). These differences were much larger (e.g., 6.34 vs. 4.70 when comparing atheists to Evangelical Protestants) prior to adjusting for the covariates in the model (i.e., education, household income, and conservative political orientation) but remained even after using adjusted means. Table 3 depicts the adjusted means and standard error (i.e., adjusted for covariates), as well as the unadjusted means, standard deviation, and corresponding number of participants in each group.

**Table 3.** Adjusted and unadjusted means for support among religious and non-religious groups, including standard deviations and sample size.

| Group | *Adjusted* |  | *Unadjusted* |  | *N* |
| --- | --- | --- | --- | --- | --- |
|  | *M* | *SE* | *M* | *SD* |  |
| Evangelical Protestant | 5.18 | 0.07 | 4.70 | 2.04 | 528 |
| Spiritual | 5.22 | 0.08 | 5.35 | 1.85 | 396 |
| Black Protestant | 5.29 | 0.07 | 5.19 | 1.65 | 410 |
| Catholic | 5.46 | 0.05 | 5.37 | 1.64 | 789 |
| Muslim | 5.49 | 0.08 | 5.52 | 1.56 | 329 |
| Mainline Protestant | 5.50 | 0.07 | 5.32 | 1.75 | 523 |
| Jewish | 5.56 | 0.08 | 5.65 | 1.58 | 364 |
| Agnostic | 5.74 | 0.07 | 6.04 | 1.32 | 498 |
| Atheist | 5.91 | 0.07 | 6.34 | 1.18 | 486 |
| TOTAL |  |  |  |  | 4323 |

Note. Covariates factored in for the adjusted means are evaluated at the following values: education = 4.28, household income = 3.22, conservative political orientation = 4.43

### Analysis 2: ANCOVA comparing concerns about mRNA vaccines among religious and non-religious groups

The ANCOVA results indicated statistically significant small sized differences (partial *η*^2^ = .06) across the religious and non-religious groups in terms of concerns about mRNA vaccines, *F (*8,4439) = 36.83, *p* < .001). Pairwise comparison between groups suggested that the Muslim group had a significantly higher mean for concerns about the use of mRNA vaccines compared to all other groups (Δ*M* ranged from 0.37 to 1.61, with all *p*s < .05), except for the Black Protestant group (Δ*M* = 0.36, *p* > .05). As in the prior model, group differences were much larger (e.g., 2.28 vs 4.09 when comparing atheists to Muslims) prior to adjusting for the covariate in the model (i.e., conservative political orientation) but remained even after using adjusted means. Table 4 depicts the adjusted means and standard error (i.e., adjusted for covariates), as well as the unadjusted means, standard deviation, and corresponding number of participants in each group.

**Table 4.** Adjusted and unadjusted means for concerns among religious and non-religious groups, including standard deviations and sample size.

| Group | Adjusted |  | Unadjusted |  | N |
| --- | --- | --- | --- | --- | --- |
|  | M | SE | M | SD |  |
| Atheist | 2.82 | 0.08 | 2.28 | 1.47 | 498 |
| Agnostic | 3.23 | 0.07 | 2.84 | 1.65 | 510 |
| Jewish | 3.50 | 0.08 | 3.48 | 1.78 | 381 |
| Mainline Protestant | 3.58 | 0.07 | 3.82 | 1.84 | 541 |
| Catholic | 3.85 | 0.06 | 4.01 | 1.86 | 807 |
| Spiritual | 3.92 | 0.08 | 3.62 | 1.89 | 411 |
| Evangelical Protestant | 4.06 | 0.07 | 4.62 | 1.84 | 536 |
| Black Protestant | 4.07 | 0.08 | 4.09 | 1.65 | 419 |
| Muslim | 4.43 | 0.09 | 4.52 | 1.70 | 336 |
| TOTAL |  |  |  |  | 4439 |

*Note.* For the adjusted mean, conservative political orientation was factored in at the 4.43, on a scale of 1 (“extremely liberal”) to 9 “(extremely conservative)”

### Analysis 3: What religious beliefs and practices predict support for mRNA vaccines?

The eight predictors selected for inclusion were entered into a hierarchical regression using the model building (training) sample. We then evaluated the reliability of the model in the reliability testing (holdout) sample. The shrinkage on cross-validation was .018, which was smaller than .10 suggested threshold, supporting the conclusion that this was a reliable model.(58)

As seen in Table 5, model had a moderate effect size (Δ*R^2^*= .13).(62) The religious variables that remained statistically significant predictors (*p*s < .0023) were: more spiritual community support for community health (*β* = 0.23, *p* < .001), higher acceptance of evolution (*β* = 0.19, *p* < .001), higher frequency of attendance in religious or spiritual activities (*β* = 0.12, *p* < .001), lower fundamentalist religious beliefs (*β* = -0.08, *p* = .001), and more spiritual community support for liberal reproductive and end of life views (*β* = 0.07, *p* < .001). Additionally, two covariates and one demographic variable significantly predicted support: lower conservative political orientation (*β* = -0.19, *p* < .001), less distrust towards the healthcare system (*β* = -0.16, *p* < .001), and higher household income (*β* = 0.11, *p* < .001).

**Table 5.**
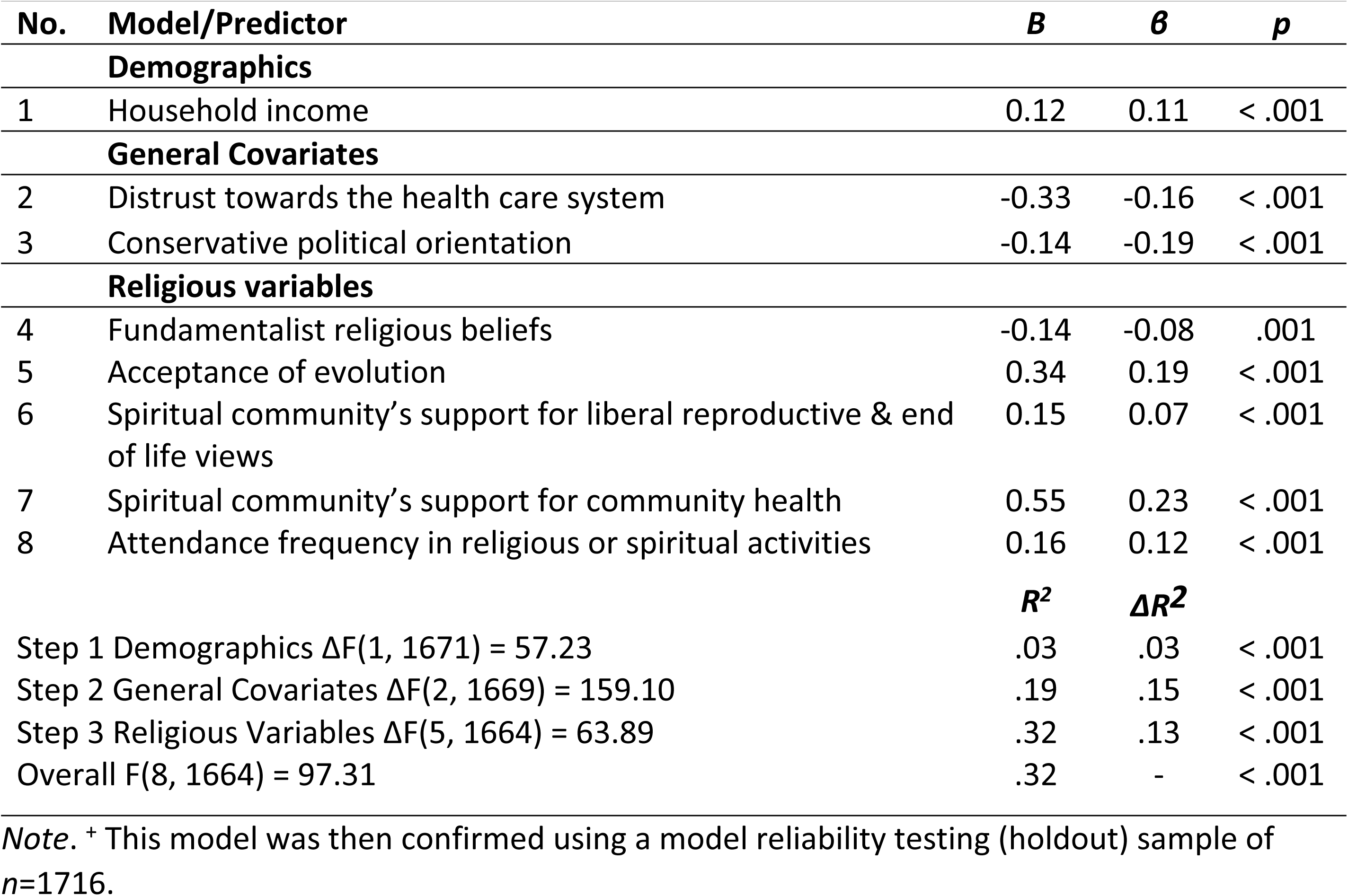
Final model for predicting support for the use of mRNA vaccine in a sample of *n* = 1715^+^.

### Analysis 4: What religious beliefs and practices predict concerns about mRNA vaccines?

Similar to the prior regression, we first entered the six selected predictors into a hierarchical regression using the model building (training) sample, then evaluated the reliability of the model in the model reliability testing (holdout) sample. The shrinkage on cross-validation was .050, which was smaller than .10 suggested threshold(58), supporting the conclusion that this was a reliable model.

As seen in Table 6, model had a small effect size (Δ*R^2^*= .06).(62) The religious variables that remained statistically significant predictors (*p*s < .0023) were: less spiritual community support for community health (*β* = -0.15, *p* < .001), stronger beliefs about God in the body (*β* = 0.14, *p* < .001), and more fundamentalist religious beliefs (*β* = 0.13, *p* < .001). The strongest predictors of concerns were some of the covariates: more distrust toward the healthcare system (*β* = 0.32, *p* < .001) and higher conservative political orientation (*β* = 0.23, *p* < .001). Lower genetic knowledge (*β* = -0.11, *p* < .001) was also a significant predictor.

**Table 6.**
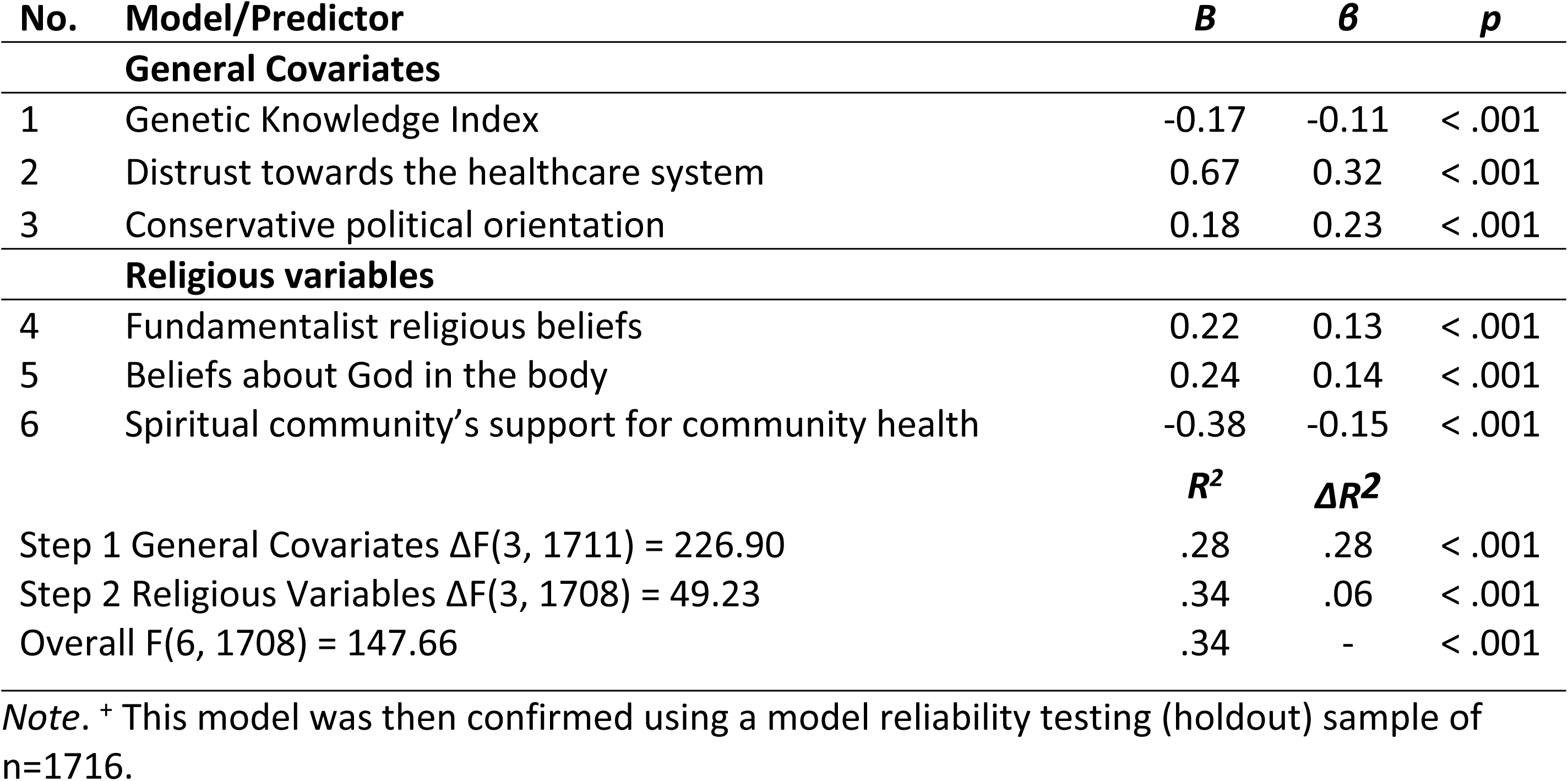
Final model for predicting concerns about the use of mRNA vaccine in a sample of *n* = 1715^+^.

## Discussion

We found that participants across religious groups generally supported mRNA vaccines and had modest levels of concerns, however there were significant differences between the religious groups. The strongest predictor of support for mRNA vaccines was their spiritual community’s support for community health, a more liberal political orientation, and higher acceptance of evolution. Regarding concerns with mRNA vaccines, the covariates of distrust toward the healthcare system and more conservative political orientation were the strongest predictors.

While we observed support for mRNA vaccines across the sample, there were significant differences between the religious groups. In particular, Evangelical Protestant participants had significantly lower support for mRNA vaccines than all other religious groups, while atheist participants had the most support. Similarly, Muslim participants reported the highest level of concerns about mRNA vaccines, while atheist participants reported the lowest concerns. These findings align with past research suggesting that some religious groups, such as Protestant and spiritual individuals, have lower support for vaccines.(23, 24, 64)

For both support and concerns, each religious group mean had a relatively large standard deviation, indicating considerable variability within each group. Given the wide range of attitudes within each group, these findings suggest that religious affiliation alone is not necessarily a strong predictor of one’s level of support or concerns about mRNA vaccines.

Among those who identified with a religion, the strongest predictor of support for mRNA vaccines was their spiritual community’s support for community health. This indicates that religious individuals place significant value on their community’s perspective regarding health-related matters. Religious individuals tend to be higher in group-focused “binding” moral values, which support and enable group cohesion.(15, 65) This finding supports the notion that their spiritual community, with which religious individuals feel connected, impacts their attitudes toward mRNA vaccines. This is noteworthy, considering vaccination decisions are often perceived as individual health decisions. Public health efforts to build trust among religious groups should consider engaging spiritual communities in order to increase mRNA vaccine acceptance and uptake.

The second strongest predictor of support was more acceptance of evolution. Views on evolution may reflect an individual’s acceptance of science and beliefs in creationism, thus, higher levels of acceptance of evolution being associated with more support for mRNA vaccines is understandable.(50) This aligns with prior research indicating these two variables (i.e., their spiritual community’s support for community health and acceptance of evolution) predict support for genetic and genomic healthcare more broadly.(7) Other significant predictors were more liberal political orientation, lower levels of distrust toward the healthcare system, more frequently attending religious activities, and lower fundamentalist religious beliefs.

Regarding concerns about mRNA vaccines, distrust toward the healthcare system was the strongest predictor, followed by more conservative political orientation. Both of these covariates were stronger predictors of concerns than they were of support for mRNA vaccines. They were also stronger predictors of concerns than any of the religious or demographic variables in the study. This is in contrast with previous research findings that religious variables were stronger predictors of attitudes genomic and genetic healthcare than political orientation, distrust, or demographics.(7)

Other significant predictors of concerns for mRNA vaccines included lower genetic knowledge, more fundamentalist religious beliefs, beliefs about God in the body, and lower beliefs that your spiritual community supports community health. This makes good sense, as higher fundamentalist religious beliefs may lead an individual to prioritize faith-based healing such as prayer or other spiritual practices over medical interventions.(8) Higher beliefs of God in the body, that one’s body is a gift from God or is created in God’s image, may lead individuals to view bodily interventions like vaccines as unnatural or as disturbing the purity of the body.(8) And similar to earlier, it makes sense that lower levels of their spiritual community’s support for community health would predict concerns with mRNA vaccines, as religious individuals are likely to value on their community’s perspective regarding vaccines.

The finding that political orientation and distrust in the healthcare system were predictors of both support and concerns about mRNA vaccines underscores the culture war around mRNA vaccines within society and distrust of science and healthcare.(1, 2) These data were collected in 2023, and the politicization of mRNA vaccines has persisted since that time. These findings indicate a need for not just for engaging religious and spiritual communities in trust-building efforts, but also the need to target and try to reduce the politicization of mRNA vaccines. These findings are also interesting in the sense that these may be instances of “gut reactions” type of thinking involved in moral decision-making.(16, 17) If participants had been asked directly why they have concerns about mRNA vaccines, they may have provided various religious beliefs and practices as reasons for their concerns. In reality, at least according to the current study’s results, their political leanings and distrust of healthcare were stronger factors in their concerns than any of the religious beliefs and practices in the study, lending support to the idea that religious concerns about vaccines may actually reflect personal beliefs rather than theological objections.(14)

Last, the dispersion of scores for support vs. concerns of mRNA vaccines were very different. The distribution for support was skewed such that a majority of participants indicated high levels of support for mRNA vaccines. However, the dispersion for concerns was much different, with the distribution spanning the full range of responses. These findings suggest that support and concerns about mRNA vaccines are separate constructs, and do not likely represent opposite sides of the same spectrum. When asking about overall support, it’s possible that participants provided a socially desirable response indicating general support, or that in theory they support mRNA vaccines. However, when asked about concerns, their responses indicate a wide variety of responses, perhaps reflective of the wide variety of views on mRNA vaccines in society. The concerns items were also more specific than the support item, and participants may support mRNA vaccines, but still have considerable concerns that may stop them from engaging with them.

This secondary analysis had several strengths. The dataset that was used was very large and contained large numbers of many major religious groups in the U.S. This allowed for comparisons of these major religious groups on their attitudes toward mRNA vaccines, whereas most of the literature in this area has been theoretical or conducted with specific religious communities.(8, 11, 12, 27-32) Additionally, the dataset contained a large number of religious beliefs and practices, which allowed for analyses to determine which of these beliefs and practices were most influential in regard to attitudes toward mRNA vaccines. Finally, the dataset was largely representative of the U.S. in terms of age, gender, and race, which aids in the generalizability of these findings.

There were some limitations to the study. One limitation was that the study was a secondary analysis of an existing dataset. Thus, research questions were limited to exploring the variables present in the dataset. While the dataset used for this analysis contained large enough numbers to compare several major religious groups in the United States, it did not allow for comparison or insights into smaller religious groups. For example, from this dataset, there were too few Buddhist, Hindu, Mormon, or other common religious groups present in the U.S. to allow for inclusion in the analysis.

In conclusion, the study identified differences in support and concerns with mRNA vaccines among several religious groups in the U.S., finding that within group differences were often larger than between group differences. Furthermore, we aimed to determine which religious beliefs and practices predicted support and concerns with mRNA vaccines and found that their spiritual community’s support for community health was the strongest religious factor impacting their attitudes, while political orientation and distrust in the healthcare system were also important factors. These findings add to the literature on the culture wars surrounding mRNA vaccines, and can perhaps aid in future efforts to build trust between public health and religious communities.

## Acknowledgements

This research was supported by the National Human Genome Research Institute (5R01HG012830, PI: DuBois) through March 11, 2025 and supported by Washington University institutional funds thereafter. Dr. Baker was supported by the National Center For Advancing Translational Sciences of the National Institutes of Health under Award Number UL1 TR002345 (PI: Powderly). The funders had no role in study design, data collection and analysis, decision to publish, or preparation of the manuscript. The content is solely the responsibility of the authors and does not necessarily represent the official views of the National Institutes of Health. The authors declare no conflicts of interest.

## Author contributions

Conceptualization: James M. DuBois. Data curation: Erin D. Solomon and Eu Gene Chin. Formal Analysis: Eu Gene Chin and Erin D. Solomon. Funding acquisition: James M. DuBois. Methodology: James M. DuBois and Eu Gene Chin. Project administration: Kari Baldwin. Supervision: James M. DuBois. Writing—Original draft: Erin D. Solomon, Eu Gene Chin, James M. DuBois. Writing—Review and editing: Erin D. Solomon, Eu Gene Chin, Kari Baldwin, Lauren L. Baker, and James M. DuBois.

## Data availability statement

The data that support the findings of this study are openly available in Open ICPSR at https://www.openicpsr.org/openicpsr/project/247241/version/V1/view

